# Excess risk of Covid-19 infection and mental distress in healthcare workers during successive pandemic waves. Analysis of matched cohorts of healthcare workers and community referents in Alberta, Canada

**DOI:** 10.1101/2023.09.12.23295439

**Authors:** Jean-Michel Galarneau, France Labrèche, Quentin Durand-Moreau, Shannon Ruzycki, Anil Adisesh, Igor Burstyn, Tanis Zadunayski, Nicola Cherry

## Abstract

**Objectives:** To investigate changes in risk of infection and mental distress in healthcare workers (HCWs) relative to the community as the Covid-19 pandemic progressed.

**Methods:** HCWs in Alberta, Canada, recruited to an interprovincial cohort, were asked consent to link to Alberta’s administrative health database (AHDB) and to information on Covid-19 immunization and polymerase chain reaction (PCR) testing. Those consenting were matched to records of up to 5 community referents (CRs). Physician diagnoses of Covid-19 were identified in the AHDB from the start of the pandemic to 31 March 2022. Physician consultations for mental health (MH) conditions (anxiety, stress/adjustment reaction, depressive) were identified from 1 April 2017 to 31 March 2022. Relative risk for HCWs was estimated for each condition, overall and for successive infection waves.

**Results:** 80% (3050/3812) of HCWs consented to be linked to the AHDB: 97% (2959/3050) were matched to 14546 CRs. HCWs were at greater risk of Covid-19 overall, with first infection defined either from PCR tests or physician records They were also at increased risk for each of the three MH diagnoses. In analyses adjusted for confounding, risk of Covid-19 infection was higher than CRs early in the pandemic and during the fifth (Omicron) wave. The excess risk of stress/adjustment reactions and depressive conditions increased with successive waves during the epidemic, peaking in the 4^th^ wave.

**Conclusion:** Administrative health data, although not a complete reflection of infection or MH, contributed to an understanding of changing risk over time, with excess risk continuing late in the pandemic

## Introduction

Healthcare workers (HCWs) were at increased risk of Covid-19 infection early in the Covid-19 pandemic [1] [2], but there is some evidence that this excess risk decreased as the pandemic moved forward [3] [4] [5] [6]. It is unclear whether this reflected improved workplace infection prevention and control practices, vaccination or a combination of these factors. Further, although there is little doubt that living and working through the pandemic was stressful for the population as a whole [7] and for HCWs [8] [9], this again seems to have reduced as the pandemic progressed, although results from longitudinal studies of HCWs are inconsistent [10]. There are rather few studies directly comparing infection rates [11] or mental health (MH) [12] [13] in HCWs with those in the general population. The study reported here uses administrative health records, including those for immunization and the results of polymerase chain reaction (PCR) testing, to assess whether HCWs were indeed at greater risk than the general population in Alberta of Covid-19 infection and of adverse MH outcomes during the Covid-19 pandemic and to chart how these risks evolved as the pandemic progressed.

## Methods

HCWs were recruited from four Canadian provinces (Alberta, British Columbia, Ontario and Quebec) during the early months of the pandemic and followed up through periodic questionnaires to the summer of 2022 [14]. Participants from Alberta were asked at recruitment for consent to match their individual records to the Alberta Administrative Health Database (AHDB). In Alberta (as across Canada) health care is free at the point of service but for physicians to be paid for a service they must enter at least one diagnosis which is recorded in the AHDB. With individual consent, the administrative database containing such records can be made available for research. As the pandemic progressed, participants were also asked for consent to be linked to Covid-19 immunization records maintained by the provinces and for results of all PCR testing for the SARS-CoV-2 virus. With research ethics board agreement, Alberta Health attempted to match the consenting HCWs to 5 anonymized community referents (CRs) on sex, age (+/- 3 years), geographic location in Alberta and number of physician claims from April 1, 2019 - March 31, 2020. Referents were alive and resident in the province on March 31, 2020 as determined by the Alberta Health population registry. Diagnostic data were extracted from inpatient care (up to 25 codes per episode), ambulatory care (up to 10 codes) and practitioner physician claims (up to 3 codes) for specified conditions, including mental ill-health and Covid-19. Alberta Health linked both HCWs and matched CRs to records of each PCR test (negative or positive) and vaccination (date and type) received. Data on immunizations and PCR tests were extracted from the start of the pandemic in Alberta (taken as 6 March 2020) to 31 March 2022. Data on the selected physician diagnoses were made available from 1 April 2017 to 31 March 2022, giving records for 35 months and 5 days before the start of the pandemic and for 24 months and 26 days during the pandemic.

For the analysis reported here, incident Covid-19 infection was examined using two criteria:

I. the first date on which a participant had received a positive PCR test and
II. the first date of a physician consultation at which the specific code for Covid-19 infection (ICD-9 079.82: ICD-10 U07.1) had been recorded.

Neither definition was held to be a comprehensive or unbiased reflection of infection in the community.

Three MH conditions (anxiety disorder, stress reaction or adjustment disorder and depressive disorder) were identified from physician records, as follows:

Anxiety disorders: ICD-9 300, 300.1, 300.2, 300.3, 300.8, 300.9; ICD-10 F40, F41, F42, F44, F45 F48.

Stress and adjustment reactions: ICD-9 308, 309; ICD-10 F43.

Depressive disorders: ICD-9 300.4 311; ICD-10 F32 F33 F34.1

Episodes were classified by the first date recorded as either before or during the pandemic. If a condition was recorded by a physician both before and after March 6^th^ 2020, it was included in both periods.

Potential covariates of interest for infection included number and dates of immunization against Covid-19, the total number of PCR tests recorded and the stage of the pandemic as reflected in infection waves in Alberta. These, rounded to the end of the previous month, were taken as: wave 1: March-June 2020 (4 months), wave 2: July 2020-February 2021 (8 months), wave 3: March-June 2021 (4 months), wave 4: July -October 2021 (4 months), wave 5 (Omicron): November 2021-March 2022 (5 months). For diagnosis of Covid-19 from results of PCR testing, the total number of tests recorded (positive or negative) was also considered. Those with no tests could not have a positive result while those with multiple tests (for whatever reason) were more likely to have an adventitious positive result, even if asymptomatic. For MH the covariates were physician records of the same MH condition in the 35 months pre-pandemic.

### Statistical methods

Odds ratios of MH diagnoses were estimated by conditional logistic regression, allowing for matching of HCWs to CRs and, in the MH analysis, for diagnoses recorded before the pandemic. To examine the evolution of risk during the pandemic, time to first Covid-19 diagnosis was examined for HCWs and CRs in a multilevel piecewise exponential model, nesting participants within matching groups [15]. The time variable was months within each wave and the hazard was assumed constant within each wave. The analysis controlled for the number of vaccines received at the time of the event (Covid-19 diagnosis) and in the case of PCR positivity, the number of tests recorded in that month. For the MH analysis, a similar approach was adopted where individual slopes were fitted within wave for the effects of being a HCW on incidence of each MH condition within the wave, with nesting of participants within matching group. The analysis was carried out in Stata 18 (StataCorp. 2023. Stata Statistical Software: Release 18. College Station, TX: StataCorp LLC)

## Results

Of 3812 Alberta HCWs who consented to join the cohort, 80% (3050/3812) agreed to be matched to the Alberta AHDB. 97% (2959/3050) were identified in the database and matched to at least 3 community referents. The analysis reported here was based on 2959 HCWs and 14,546 CRs.

Among the HCWs 476 (16.1%) were physicians, 2353 (79.5%) were registered nurses, 58 (2.0%) licensed practical nurses and 72 (2.4%) health care aides. Most were female (87.5%,2590/2959). The median age at recruitment was 44 years.

HCWs were more likely to have a Covid-19 infection, whether identified by a PCR test (20% of HCWs had at least one positive PCR test compared to 12% of CRs) or by medical records (HCWs 32%; CRs 26%): 44% of HCWs and 33% of CRs were identified by one or other case definition (Table 1). Twenty HCWs and 32 CRs had a second positive PCR test. Repeat infection could not be confidently identified from physician diagnoses and only first cases have been considered here.

**Table 1.** First episode of Covid-19 and mental health conditions in healthcare workers and community referents.

| Health condition | Healthcare workers |  | Community referents |  | Both |  | Odds ratio* | 95% CI | P-value |
| --- | --- | --- | --- | --- | --- | --- | --- | --- | --- |
|  | N | % | N | % | N | % |  |  |  |
| **Covid-19** |  |  |  |  |  |  |  |  |  |
| Any positive PCR test report | 599 | 20.2 | 1689 | 11.6 | 2288 | 13.1 | 1.96 | 1.76 to 2.17 | <0.001 |
| Any Covid-19 medical report | 941 | 31.8 | 3831 | 26.3 | 4772 | 27.3 | 1.33 | 1.21 to 1.45 | <0.001 |
| Either report | 1303 | 44.0 | 4826 | 33.2 | 6129 | 35.0 | 1.62 | 1.49 to 1.75 | <0.001 |
| **Any medical report of** |  |  |  |  |  |  |  |  |  |
| *Anxiety condition* |  |  |  |  |  |  |  |  |  |
| Before pandemic | 809 | 27.3 | 4108 | 28.2 | 4917 | 28.1 | 0.96 | 0.87 to 1.05 | 0.345 |
| During pandemic | 866 | 29.3 | 3801 | 26.1 | 4667 | 26.7 | 1.19 | 1.08 to 1.30 | <0.001 |
| Adjusted for before pandemic |  |  |  |  |  |  | 1.25 | 1.13 to 1.38 | <0.001 |
| *Stress/adjustment reaction* |  |  |  |  |  |  |  |  |  |
| Before pandemic | 381 | 12.9 | 1802 | 12.4 | 2183 | 12.5 | 1.05 | 0.93 to 1.18 | 0.438 |
| During pandemic | 445 | 15.0 | 1556 | 10.7 | 2001 | 11.4 | 1.50 | 1.33 to 1.68 | <0.001 |
| Adjusted for before pandemic |  |  |  |  |  |  | 1.52 | 1.35 to 1.71 | <0.001 |
| *Depressive condition* |  |  |  |  |  |  |  |  |  |
| Before pandemic | 647 | 21.9 | 2856 | 19.6 | 3503 | 20.0 | 1.16 | 1.05 to 1.28 | 0.004 |
| During pandemic | 652 | 22.0 | 2490 | 17.1 | 3142 | 17.9 | 1.39 | 1.26 to 1.54 | <0.001 |
| Adjusted for before pandemic |  |  |  |  |  |  | 1.39 | 1.24 to 1.55 | <0.001 |
| **N** | 2959 |  | 14546 |  | 17505 |  |  |  |  |

\*From conditional logistic regression

Examination of mental health diagnoses showed that HCWs were more likely than CRs to have a physician record of an anxiety, stress/adjustment reaction or depressive condition during the pandemic. Before the pandemic there was no difference in the proportion of HCWs and CRs with a record of anxiety or stress/adjustment reaction but HCWs were somewhat more likely to have a record of a depressive condition. The risk estimates (odds ratios) given in Table 1 show an increased risk for HCWs of all three MH diagnoses during the pandemic, having adjusted for reports of the same condition before the pandemic.

The distribution of Covid-19 cases identified by each criterion is shown in Figure 1, which also indicates the infection waves. Cases identified by PCR tests were predominantly in the later stages of the pandemic, while those from physician records peaked in the first wave. HCWs had more PCR tests, positive or negative (median 3.0) than CRs (median 1.0) during the course of the pandemic, with the highest mean rate of tests/month in wave 2 (Table 2). In both HCWs and CRs there was a marked increase in the proportion of tests that were positive in wave 5. Among those tested, the proportion of tests that were positive was somewhat lower for HCWs. In each wave after the first (when vaccines had not yet been introduced), HCWs had received more vaccination doses than CRs.

**Table 2.** First episode of Covid-19, polymerase chain reaction (PCR) tests and vaccine shots by infection wave: healthcare workers (HCW, N=2959) and community referents (CR, N=14546).

|  | Wave |  |  |  |  |  |  |  |  |  |
| --- | --- | --- | --- | --- | --- | --- | --- | --- | --- | --- |
| Covid-19 medical report | 1<br>March-June 2020 |  | 2<br>July 2020-February 2021 |  | 3<br>March-June 2021 |  | 4<br>July-October 2021 |  | 5<br>November 2021 - March 2022 |  |
|  | N | % | N | % | N | % | N | % | N | % |
| HCW | 457 | 15.4 | 255 | 8.6 | 107 | 3.6 | 28 | 0.9 | 94 | 3.2 |
| CR | 1701 | 11.7 | 996 | 6.8 | 548 | 3.8 | 222 | 1.5 | 364 | 2.5 |
| All | 2158 | 12.3 | 1251 | 7.1 | 655 | 3.7 | 250 | 1.4 | 458 | 2.6 |
| **Positive PCR** |  |  |  |  |  |  |  |  |  |  |
| HCW | 7 | 0.2 | 104 | 3.5 | 22 | 0.7 | 44 | 1.5 | 442 | 14.9 |
| CR | 12 | 0.1 | 370 | 2.5 | 278 | 1.9 | 275 | 1.9 | 785 | 5.4 |
| All | 19 | 0.1 | 474 | 2.7 | 300 | 1.7 | 319 | 1.8 | 1227 | 7.0 |
| **Any PCR test** |  |  |  |  |  |  |  |  |  |  |
| HCW | 973 | 32.9 | 1984 | 67.0 | 840 | 28.4 | 986 | 33.3 | 1353 | 45.7 |
| CR | 1605 | 11.0 | 5768 | 39.7 | 2688 | 18.5 | 2674 | 18.4 | 2599 | 17.9 |
| All | 2578 | 14.7 | 7752 | 44.3 | 3528 | 20.2 | 3660 | 20.9 | 3952 | 22.6 |
| **Two or more vaccine shots** |  |  |  |  |  |  |  |  |  |  |
| HCW | 0 | 0.0 | 1471 | 49.7 | 2675 | 90.4 | 2856 | 96.5 | 2870 | 97.0 |
| CR | 0 | 0.0 | 510 | 3.5 | 6855 | 47.1 | 11390 | 78.3 | 11946 | 82.1 |
| All | 0 | 0.0 | 1981 | 11.3 | 9530 | 54.4 | 14246 | 81.4 | 14816 | 84.6 |
|  | Mean | SD | Mean | SD | Mean | SD | Mean | SD | Mean | SD |
| **Total PCR tests** |  |  |  |  |  |  |  |  |  |  |
| HCW | 0.39 | 0.63 | 1.80 | 2.56 | 0.45 | 0.91 | 0.46 | 0.79 | 0.70 | 1.00 |
| CR | 0.12 | 0.38 | 0.76 | 1.52 | 0.28 | 0.88 | 0.26 | 0.80 | 0.24 | 0.73 |
| All | 0.17 | 0.44 | 0.94 | 1.78 | 0.30 | 0.89 | 0.29 | 0.81 | 0.32 | 0.80 |
| **Proportion of PCR tests positive** |  |  |  |  |  |  |  |  |  |  |
| HCW | 0.01 | 0.07 | 0.03 | 0.14 | 0.02 | 0.11 | 0.04 | 0.18 | 0.24 | 0.39 |
| CR | 0.01 | 0.08 | 0.04 | 0.18 | 0.09 | 0.27 | 0.09 | 0.27 | 0.26 | 0.41 |
| All | 0.01 | 0.08 | 0.04 | 0.17 | 0.07 | 0.24 | 0.08 | 0.25 | 0.25 | 0.40 |
| **Vaccine shots** |  |  |  |  |  |  |  |  |  |  |
| HCW | 0.00 | 0.00 | 1.07 | 0.96 | 1.85 | 0.48 | 2.00 | 0.43 | 2.79 | 0.60 |
| CR | 0.00 | 0.00 | 0.08 | 0.38 | 1.19 | 0.84 | 1.66 | 0.84 | 2.15 | 1.07 |
| All | 0.00 | 0.00 | 0.25 | 0.64 | 1.31 | 0.83 | 1.72 | 0.79 | 2.26 | 1.03 |

**Figure 1.**
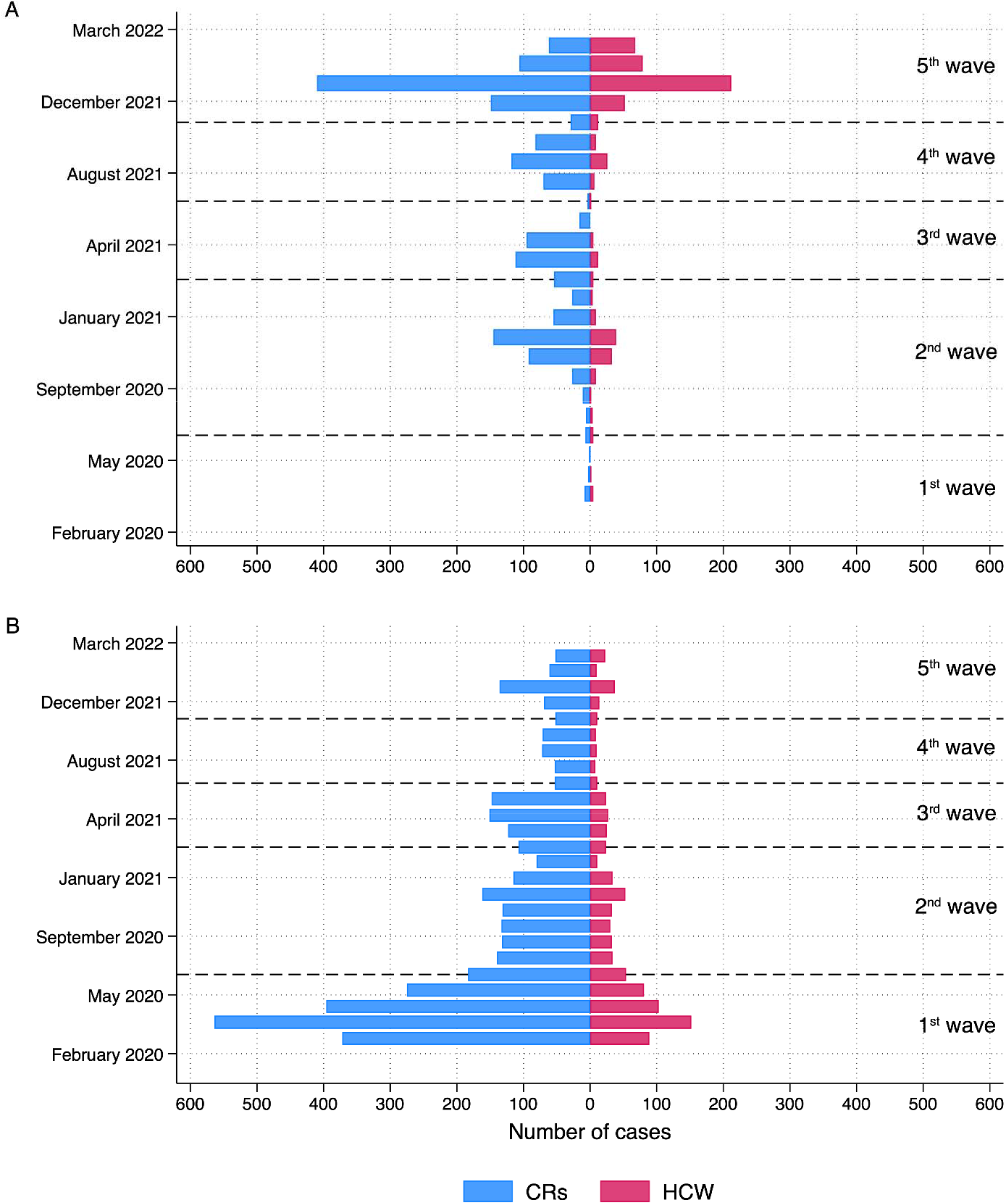
Distribution of cases by date and wave. A) Cases from PCR test, B) cases from physician records.

Table 3 gives multivariable analyses for Covid-19, computed within wave and adjusted for the number of vaccinations received and the number of PCR tests in the month of infection. Risk (hazard ratio) to HCWs, relative to CRs, of Covid-19 defined from physician records decreased with successive infection waves until the 5^th^ (Omicron) wave. A similar pattern was evident for cases defined by PCR test, with a parallel, but greater, increase in wave 5. Risk decreased with vaccination where the criterion was a positive PCR test. The relation of immunization to cases in physician records showed a contrary pattern.

**Table 3.** Hazard of Covid-19 (healthcare workers relative to community referents) by infection wave adjusted for covariates.

|  | Covid-19 |  |  |  |  |  |
| --- | --- | --- | --- | --- | --- | --- |
|  | From physician record |  |  | From PCR test |  |  |
|  | HR* | 95% CI | p= | HR* | 95% CI | p= |
| **Infection wave** |  |  |  |  |  |  |
| **1: March-June 2020** | 1.30 | 1.17 to 1.45 | <0.001 | 1.67 | 0.60 to 4.61 | 0.323 |
| **2: July 2020-Feb 2021** | 1.31 | 1.15 to 1.50 | <0.001 | 0.51 | 0.37 to 0.71 | <0.001 |
| **3: March-June 2021** | 0.93 | 0.75 to 1.17 | 0.553 | 0.41 | 0.24 to 0.69 | 0.001 |
| **4: July-October 2021** | 0.78 | 0.55 to 1.10 | 0.158 | 0.77 | 0.52 to 1.14 | 0.196 |
| **5: Nov 2021-March 2022** | 1.31 | 1.03 to 1.66 | 0.026 | 2.45 | 2.00 to 3.01 | <0.001 |
| **Numbers of vaccine doses** |  |  |  |  |  |  |
| **0** | 1.00 | – | – | 1.00 | – |  |
| **1** | 1.48 | 1.27 to 1.71 | <0.001 | 0.99 | 0.89 to 1.24 | 0.961 |
| **2** | 1.27 | 1.10 to 1.46 | 0.001 | 0.55 | 0.45 to 0.68 | <0.001 |
| **3** | 1.31 | 1.05 to 1.62 | 0.016 | 0.74 | 0.58 to 0.94 | 0.016 |
| **4** | 3.80 | 1.55 to 9.29 | 0.003 | 0.32 | 0.06 to 1.69 | 0.178 |
| **Number of PCR tests** | – | – | – | 6.45 | 6.09 to 6.83 | <0.001 |
| **N observations** | 102145 |  |  | 152140 |  |  |
| **Individuals** | 17505 |  |  | 17505 |  |  |
| **Groups** | 2959 |  |  | 2959 |  |  |
| **Number of failures** | 4772 |  |  | 2288 |  |  |

\*Hazard ratio of health care workers relative to community referents from multiple multilevel piecewise exponential proportional hazards regression

The evolution of MH risk by wave is shown in Table 4. The incidence within each wave is given in the top half of the table, with anxiety being, overall the most reported condition and stress/adjustment reaction the least. Wave 2, covering 8 months, has higher absolute number of reports than the waves covering shorter periods. The odds ratio for HCWs in each wave is given in the lower half of Table 4. There was no excess of anxiety for HCWs during the first wave of the pandemic but small increased risks of conditions coded as stress, adjustment reaction or depressive conditions were present at this point. The excess in anxiety conditions, though present in all subsequent waves, was less evident than those for depressive conditions or stress/adjustment reactions, which showed an increasing trend as the pandemic progressed, peaking at wave 4.

**Table 4.** Physician reports of anxiety, stress/adjustment reaction and depressive conditions by infection wave for healthcare workers (HCW) and community referents (CR).

|  | Wave |  |  |  |  |  |  |  |  |  |  |
| --- | --- | --- | --- | --- | --- | --- | --- | --- | --- | --- | --- |
|  | 1<br>March-June 2020 |  | 2<br>July 2020-February 2021 |  | 3<br>March-June 2021 |  | 4<br>July-October 2021 |  | 5<br>November 2021-March 2022 |  | N |
| Anxiety | N | % | N | % | N | % | N | % | N | % |  |
| HCW | 297 | 10.0 | 446 | 15.1 | 325 | 11.0 | 278 | 9.4 | 348 | 11.8 | 2959 |
| CR | 1431 | 9.8 | 1950 | 13.4 | 1514 | 10.4 | 1245 | 8.6 | 1524 | 10.5 | 14546 |
| All | 1728 | 9.9 | 2396 | 13.7 | 1839 | 10.5 | 1523 | 8.7 | 1872 | 10.7 | 17505 |
| Stress/adjustment reaction |  |  |  |  |  |  |  |  |  |  |  |
| HCW | 102 | 3.4 | 177 | 6.0 | 144 | 4.9 | 120 | 4.1 | 141 | 4.8 | 2959 |
| CR | 392 | 2.7 | 650 | 4.5 | 465 | 3.2 | 352 | 2.4 | 429 | 2.9 | 14546 |
| All | 494 | 2.8 | 827 | 4.7 | 609 | 3.5 | 472 | 2.7 | 570 | 3.3 | 17505 |
| Depression |  |  |  |  |  |  |  |  |  |  |  |
| HCW | 234 | 7.9 | 360 | 12.2 | 264 | 8.9 | 246 | 8.3 | 285 | 9.6 | 2959 |
| CR | 964 | 6.6 | 1329 | 9.1 | 1011 | 7.0 | 816 | 5.6 | 1045 | 7.2 | 14546 |
| All | 1198 | 6.8 | 1689 | 9.6 | 1275 | 7.3 | 1062 | 6.1 | 1330 | 7.6 | 17505 |
| HCW risk of physician report of mental ill-health by wave, adjusted for pre-pandemic morbidity |  |  |  |  |  |  |  |  |  |  |  |
| Anxiety |  |  |  |  |  |  |  |  |  |  | 17505 |
| OR | 1.12 |  | 1.33 |  | 1.18 |  | 1.27 |  | 1.32 |  |  |
| 95% CI | 0.92 to 1.36 |  | 1.12 to 1.58 |  | 0.98 to 1.43 |  | 1.04 to 1.55 |  | 1.10 to 1.59 |  |  |
| p= | 0.263 |  | 0.001 |  | 0.081 |  | 0.017 |  | 0.003 |  |  |
| Stress/adjustment reaction |  |  |  |  |  |  |  |  |  |  | 17505 |
| OR | 1.45 |  | 1.55 |  | 1.84 |  | 2.07 |  | 1.99 |  |  |
| 95%CI | 1.11 to 1.91 |  | 1.24 to 1.94 |  | 1.44 to 2.34 |  | 1.59 to 2.69 |  | 1.55 to 2.54 |  |  |
| p= | 0.007 |  | <0.001 |  | <0.001 |  | <0.001 |  | <0.001 |  |  |
| Depression |  |  |  |  |  |  |  |  |  |  | 17505 |
| OR | 1.27 |  | 1.62 |  | 1.46 |  | 1.89 |  | 1.60 |  |  |
| 95%CI | 1.00 to 1.61 |  | 1.31 to 2.00 |  | 1.16 to 1.84 |  | 1.49 to 2.39 |  | 1.28 to 2.00 |  |  |
| p= | 0.048 |  | <0.001 |  | 0.001 |  | <0.001 |  | <0.001 |  |  |

## Discussion

Matching of healthcare workers to community referents in an administrative health database allowed comparison of cases of Covid-19 and mental ill-health more readily and economically than through recruitment and retention of an active community cohort. Although the approach had limitations (discussed below) the overall excess cases of Covid-19, using two contrasting operational definitions, and of mental ill-health adjusting for pre-pandemic morbidity, suggest that working during the pandemic was detrimental to the health of HCWs.

Both definitions of Covid-19 infections used here had limitations. Ideally, for this analysis, all participants would have received PCR tests when symptomatic, with confirmation of infection by longitudinal serology testing. More realistically it would have been helpful to know why PCR tests were done (for screening, suspected exposure or symptomatic disease) and to have had also results of rapid antigen tests once these became available. After the first wave access to PCR testing through provincial testing sites was readily available in Alberta until the late summer of 2021. With the great increase in demand for tests when the highly infective Omicron variant became dominant (wave 5), PCR testing was largely limited to those positive on a rapid antigen test, resulting in the much greater positivity rate (24% overall) for both HCWs and CRs in this period. HCWs had designated testing sites at this phase which may in part explain the higher wave 5 risk (OR=2.45) by PCR result than from physician records (OR=1.31) for the same period.

The identification of Covid-19 cases from physician records also introduced difficulties of interpretation. Many, perhaps most, individuals with a positive test for Covid-19 would not have had reason to consult a family physician or visit a walk-in clinic and indeed they would have been discouraged from doing so. Many of the consultations early in the pandemic may have been from ‘worried well’ concerned that they might have been exposed to the virus rather than from symptomatic cases: the diagnostic code recorded did not carry this degree of specificity. A further complication is that patients with ongoing health concerns after the acute period of infection may have received the same diagnostic coding and such records cannot be distinguished from repeat infections. Given these considerable limitations, finding the same pattern over time by the two approaches, with decreasing excess risk from wave 1 until the increase at wave 5, is somewhat reassuring.

A strength of the dataset is that vaccination records would have very largely been complete and accurate: very few would have been vaccinated out of province. The observation that vaccination was associated with an increased risk of infection when the case criterion was a physician record, may reflect both greater access to physicians by those who received multiple vaccinations and an unwillingness of those not vaccinated (almost entirely CRs) to seek medical attention.

The mental health data were entirely from administrative health records. These are not comprehensive as a reflection of mental ill-health as they do not include assessment of interventions from psychologists, counsellors or other health professionals who may give support. In Alberta, with a shortage of family physicians, HCWs might have easier physician access. A physician may be biased in their readiness to record a MH diagnosis for HCWs than other patients. Equally, HCWs may differ from other patients in their willingness to disclose mental distress to a treating physician. However, CRs were matched to HCWs on the number of physician contacts in the year before the pandemic and data from before the pandemic did not show differences in anxiety and stress/adjustment reactions. Assuming that patterns of physician use and willingness to disclose did not change differentially between HCWs and CRs, we can conclude that HCWs were more likely to have episodes of mental distress during the pandemic.

Further weaknesses lie in the response rate of HCWs to join the cohort (at best around 15% of those approached [14] and the unwillingness of 20% (752 participants) to give consent to data linkage at the time of recruitment. Those who took part may not be representative of HCWs in Alberta or, indeed, of HCWs elsewhere. A further potential bias may arise in comparing employed HCWs with community members matched only on gender, age, location and physician consultations in the previous year. Although such factors are likely to affect exposure to the SARS-CoV-2 virus and behavior once infected, lack of information on employment (not recorded in the AHDB) in the referents may be a source of bias if employment itself (and not simply as a HCW) is a risk for Covid-19 infection.

The results from this analysis are consistent with the observation [3] [6] that occupational risks of infection in HCWs decreased as the pandemic progressed. An excess risk in HCW relative to the community during the Omicron wave has been reported from Hong Kong, comparing infection rates in staff from a single hospital to that in the general population [16], but not more widely. Our earlier work has shown that while risk of work-related Covid-19 decreased after vaccination it did not disappear completely [6] and it seems feasible that, with a surge in infection in the population, HCWs were again at greater risk, as suggested here. The increase in risk of a diagnosis of a stress/adjustment reaction or a depressive condition from the start of the pandemic to the fourth wave (to late fall 2021) is unlikely to be due to chance. It may in part reflect a return to equanimity in the community alongside a decreasing resilience in HCWs, coping with high work demands: both factors may contribute. Previous studies have shown a deterioration in mental health in HCWs early in the pandemic [13]. In Australia a study following HCWs from May 2021 to June 2022 found a deterioration in MH [17] while one from Italy for much the same period found MH improvements [18]. A systematic review that included 18 longitudinal studies of mental health in HCWs during the pandemic concluded that 12 studies suggested deterioration over time and 6 improvement, attributing the ‘remarkable variation’ to use of different instruments for measuring MH [10].

The current study has strengths in that it uses physician records before and during the pandemic and objective testing for the SARS-CoV-2 virus. While it is appropriate to be concerned about the limitations of the data, they do provide an unusual opportunity to examine trends over time using data collected for administrative purposes. The conclusion here, as elsewhere, [2][8] is that HCWs were at risk early in the pandemic. These new data suggest that risk was not confined to the chaotic early months and may persist or recur with new waves of infection.

### Statement from the Government of Alberta

This study is based in part on data provided by Alberta Health. The interpretation and conclusions contained herein are those of the researchers and do not necessarily represent the views of the Government of Alberta. Neither the Government nor Alberta Health express any opinion in relation to this study.

## Data Availability

Data on the healthcare worker cohort are available on reasonable request. Data on the community referents was released under the condition that it was not shared further.

## Notes

### Competing Interest Statement

The authors have declared no competing interest.

### Funding Statement

The College of Physicians and Surgeons of Alberta gave seed funding for the establishment of the HCW cohort. Grant funding was obtained from the Canadian Institutes of Health Research (Funding Reference number 173209). This funding was extended by a grant from the Canadian Immunology Task Force.

### Author Declarations

The project was reviewed and approved by the Health Ethics Review Board of the University of Alberta (Pro00099700).

